# Reinfection with SARS-CoV-2: outcome, risk factors and vaccine efficacy in a Scottish cohort

**DOI:** 10.1101/2021.11.23.21266574

**Authors:** Paul M McKeigue, David A McAllister, Chris Robertson, Diane Stockton, Helen M Colhoun, for the PHS COVID-19 Epidemiology and Research Cell

## Abstract

**Background:** The objective of this study was to investigate how protection against COVID-19 conferred by previous infection is modified by vaccination.

**Methods:** In a cohort of all 152655 individuals in Scotland alive at 90 days after a positive test for SARS-CoV-2 (confirmed by cycle threshold < 30, or two tests) followed till 22 September 2021, rate ratios for reinfection were estimated with calendar time or tests as timescale.

**Findings:** Rates of detected and hospitalised reinfection with COVID-19 while unvaccinated were respectively 6.8 (95% CI 6.4 to 7.2) and 0.18 (95% CI 0.12 to 0.25) per 1000 person-months. These rates were respectively 68% and 74% lower than in a matched cohort of individuals who had not previously tested positive. Efficacy of two doses of vaccine in those with previous infection was estimated as as 84% (95 percent CI 81% to 86%) against detected reinfection and 71% (95 percent CI 29% to 88%) against hospitalised or fatal reinfection. The rate of detected reinfection after two doses of vaccine was 1.35 (95% CI 1.02 to 1.78) times higher in those vaccinated before first infection than in those unvaccinated at first infection.

**Interpretation:** The combination of natural infection and vaccination provides maximal protection against new infection with SARS-CoV-2: prior vaccination does not impair this protection.

**Funding:** No specific funding was received for this work.

**Research in context:** *Evidence before this study:* In a recent systematic review of cohort studies reported up to July 2021, the average reduction in COVID-19 infection rates in those with previous infection compared with those without evidence of previous infection was 90%. There is little information about the protective effect of previous infection against severe COVID-19, or about how the protective effects of previous infection against reinfection and severe disease are modified by vaccination.

*What this paper adds:* In unvaccinated individuals the protection against hospitalised COVID-19 conferred by previous infection is similar to that induced by vaccination. In those with previous infection, vaccination reduces the rates of reinfection and hospitalised COVID-19 by about 70%.

*Implications of all the available evidence:* The combination of natural infection and vaccination provides maximal protection against COVID-19: prior vaccination does not seriously impair this protection.

## Introduction

Although cohort studies have shown that previous SARS-Cov-2 infection confers pprotection against reinfection [1], few studies have examined how this protection is modified by vaccination before or after first infection. or by the emergence of new variants.

In the UK and most other countries those with evidence of previous SARS-CoV-2 infection are included in programmes for vaccination against COVID-19 and are not exempt from mandatory vaccine certification where this is required. This policy has been questioned as an unnecessary use of scarce vaccines, or as having an unfavourable risk-benefit ratio because adverse reactions to vaccine have been reported to be more common in the those with previous infection [2]. The evidence base for these policies depends on comparisons of rates of infection and severe disease between those with and without evidence of previous SARS-Cov-2 infection, and on estimates of the efficacy of vaccination against reinfection. The objective of this study was to investigate how the protective effect of previous infection against reinfection and against hospitalisation with COVID-19 is modified by vaccination.

## Methods

### Construction of cohort at risk of reinfection

CDC criteria for suspected reinfection are based on detection of SARS-CoV-2 RNA at least 90 days after the first detection of SARS-CoV-2 RNA, with restriction to cycle threshold (Ct) less than 33 where available [3]. All positive nucleic acid tests for SARS-CoV-2 in Scotland since 1 March 2020 were identified from the Electronic Communication of Surveillance in Scotland database (ECOSS). This database captures all tests in Pillar 1 (NHS laboratories testing those with a clinical need and workers in health or social care) and Pillar 2 (Lighthouse laboratories testing the wider population from September 2020 onwards). Only the Lighthouse laboratories report Ct values. To exclude possible false-positives, definite first infection was defined as a positive test with Ct < 30 for both N and ORF genes, or a second positive test within fourteen days of the first.

Entry date to the cohort at risk of reinfection was 90 days after first testing positive. Exit date was the earliest of date of reinfection (defined as any positive test after entry date), date of death, or end of follow-up period (22 September 2021).

### Construction of comparison cohort

To compare reinfection rates with first infection rates, we constructed a comparison cohort of individuals who had not previously tested positive for SARS-CoV-2. As all test-positive cases cases in Scotland had been sampled in the REACT-SCOT case-control study [4], for each individual in the cohort at risk of reinfection up to ten controls were available who were sampled from the general population, matched for age, sex and general practice, alive on the day that the case first tested positive, and had not tested positive by that date. As with the cohort at risk of reinfection, this comparison cohort was restricted to those who were still under observation (without having tested positive) at 90 days from the date that they were first sampled as controls. The exit date for this cohort was the earliest of date of first positive test, date of death or end of follow-up period. Those with a definite positive test who were still under observation 90 days later entered the cohort at risk of reinfection.

### Linkage to health records and occupational databases

Using the Common Healthcare Identifier (CHI) held on all Scottish health records these individuals were linked to the population register, the national vaccination database, registers of teachers and health care workers, the list of those designated as clinically extremely vulnerable (eligible for shielding), the ECOSS database of test results, a database of hospitalisations (RAPID) that is updated daily, dispensed prescriptions in primary care and death registrations as described elsewhere [4–7]. As described elsewhere individuals were clasified into three categories of clinical risk: no risk condition, designated moderate risk condition, and clinically extremely vulnerable [6].

Fatal outcome was defined as death within 28 days of testing positive or any death certified with COVID-19 as underlying cause. Hospitalisation with COVID-19 was defined as any admission within 14 days of testing positive or any positive test while in hospital. This standard definition does not distinguish those hospitalised with a primary diagnosis of COVID-19 from those hospitalised for other reasons who test positive on admission. To obtain a lower bound on the proportion of those hospitalised with but not because of COVID-19, we used Scottish Morbidity Record (SMR01) data to classify the specialty under which hospitalised cases were admitted (first episode in a continuous inpatient stay) as non-COVID (ISD Scotland specialty codes matching regular expression `^A[289DGHMR]|^[CEFHJ`) or other. The specialties classified as non-COVID include all surgical specialties and medical specialties such as oncology under which it is unlikely that patients with a primary diagnosis of COVID-19 would be admitted. SMR01 coding lags by about three months, so codes for most admissions after July 2021 were missing.

### Statistical methods

Rates of detected infection and hospitalised or fatal COVID-19 in the two cohorts were calculated separately for person-months at risk while unvaccinated, vaccinated with 1 dose and vaccinated with 2 doses. In the cohort at risk of reinfection rate ratios for hospitalised reinfection were estimated from Cox regression models, with vaccination status as a time-updated covariate. Time-invariant covariates in these models included variables previously reported as associated with severe COVID-19 in Scotland [4,6]: baseline age, sex, care home residence, occupation, and clinical risk category as a factor with three levels: no risk condition, moderate risk condition, clinically extremely vulnerable. Vaccine efficacy is 1 minus the rate ratio associated with vaccinated versus unvaccinated status [8].

For hospitalised reinfection, a Cox regression model with calendar timescale was used to estimate rate ratios as case ascertainment should be complete for this outcome. For the broader outcome of any reinfection, case ascertainment based on unscheduled testing will be incomplete and this may bias estimation of effects on infection rates where testing rates vary with the covariates of interest. Test-negative control designs do not completely overcome this [9,10]. To evaluate the sensitivity of the effect estimates to biased case ascertainment, we compared two alternative Cox regression models: a conventional Cox model with calendar timescale, and a Cox model with tests as timescale (additionally stratifying by 14-day intervals of calendar time). For the model with tests as timescale, each test from entry date to exit date (censoring at first positive test) is an observation and tests are numbered in sequence within each individual. This Cox model with tests as timescale formalizes the test-negative control design [9] as a survival analysis, as explained in the Discussion.

## Results

### Infections and hospitalisations in those with and without previous infection

Of the 532168 individuals who had tested positive since the start of the epidemic, there were 383182 who met the criteria for definite first infection: 369547 tested in Pillar 2 who had Ct < 30 and a further 16465 who had tested positive a second time within 14 days of the first positive test. Of these, 165004 were still under observation at least 90 days after first testing positive and before the end of the follow-up period on 22 September 2021, forming the cohort at risk of reinfection. Of these, 152655 had not been vaccinated at the time of first testing positive, 9725 had received one dose and 2624 had received two doses. Average follow-up of this cohort was 5 months. Figure S1 shows that the number of individuals in the cohort at risk of reinfection increased rapidly after mid-December 2020, around 90 days after Pillar 2 testing began.

The comparison cohort comprised 1177827 individuals matched for age, sex and general practice to those in the cohort at risk of reinfection, with average follow up of 5.6 months. Of these, 1132118 were unvaccinated when first sampled. The average testing rate while unvaccinated was higher in the cohort with prior infection (0.56 per month) than in the comparison cohort (0.27 per month).

In the cohort at risk of reinfection, there were 1070 detected reinfections while still unvaccinated, of which 28 were hospitalised or fatal. The corresponding rates per 1000 person-months were 6.8 (95% CI 6.4 to 7.2) for reinfections and 0.18 (95% CI 0.12 to 0.25) for hospitalisation. In the comparison cohort, there were 36488 detected first infections while still unvaccinated of which 1211 were hospitalised or fatal. The corresponding rates per 1000 person-months were 21.1 (95% CI 20.8 to 21.3) for reinfection and 0.7 (95% CI 0.66 to 0.74) for hospitalisation. This equates to a reduction in risk of 68% for detected new infection, and 74% for hospitalisation. Because the testing rate was lower in the comparison cohort than in the cohort at risk of reinfection, this comparison of detected infections is likely to underestimate the protection against new infection that is conferred by previous infection.

For the subset of hospitalised unvaccinated cases whose SMR01 records were available, the proportion admitted under non-COVID specialties was 7 of 14 in the cohort at risk of reinfection, and 47 of 421 in the comparison cohort.

### Relation of reinfection in fully vaccinated individuals to vaccination status at time of first infection

To investigate whether protection against reinfection among fully vaccinated individuals with a history of previous infection depends on whether vaccination preceded the first infection, we compared within the cohort at risk of reinfection the rates of detected reinfection after the second dose of vaccine between those who were unvaccinated at first infection and those who had received at least one dose before first infection. In these two groups there were reespectively 660 and 56 detected reinfections during follow-up. In a Cox regression model including age, sex, occupation and clinical risk category as covariates, the rate ratio for detected reinfection after the second dose of vaccine in those who had been vaccinated at least once before first infection, compared with those unvaccinated at first infection was 1.35 (95% CI 1.02 to 1.78).

### Testing rates in the cohort at risk of reinfection

Table S1 shows how crude testing rates in those who were unvaccinated at time of first infection varied with vaccination status and other covariates. This gives some idea of the likely direction of biases in estimates of the effects of these covariates on reinfection rates. Testing rates were higher in fully vaccinated than in unvaccinated or partly vaccinated time intervals, and the associations with other covariates varied by vaccination status. Testing rates were higher in women than in men, and higher in teachers and health care workers than in other occupations. In unvaccinated individuals testing rates were higher in care home residents than those living independently, higher in older than in younger age groups, and higher in those with clinical risk conditions compared with those with no risk conditions.

### Effect of vaccination and other covariates on detected reinfection

Figure 1 compares the coefficients estimated with each of the two alternative Cox regression models for detected reinfection. Table S2 tabulates the covariate distributions and coefficients.

**Figure 1:**
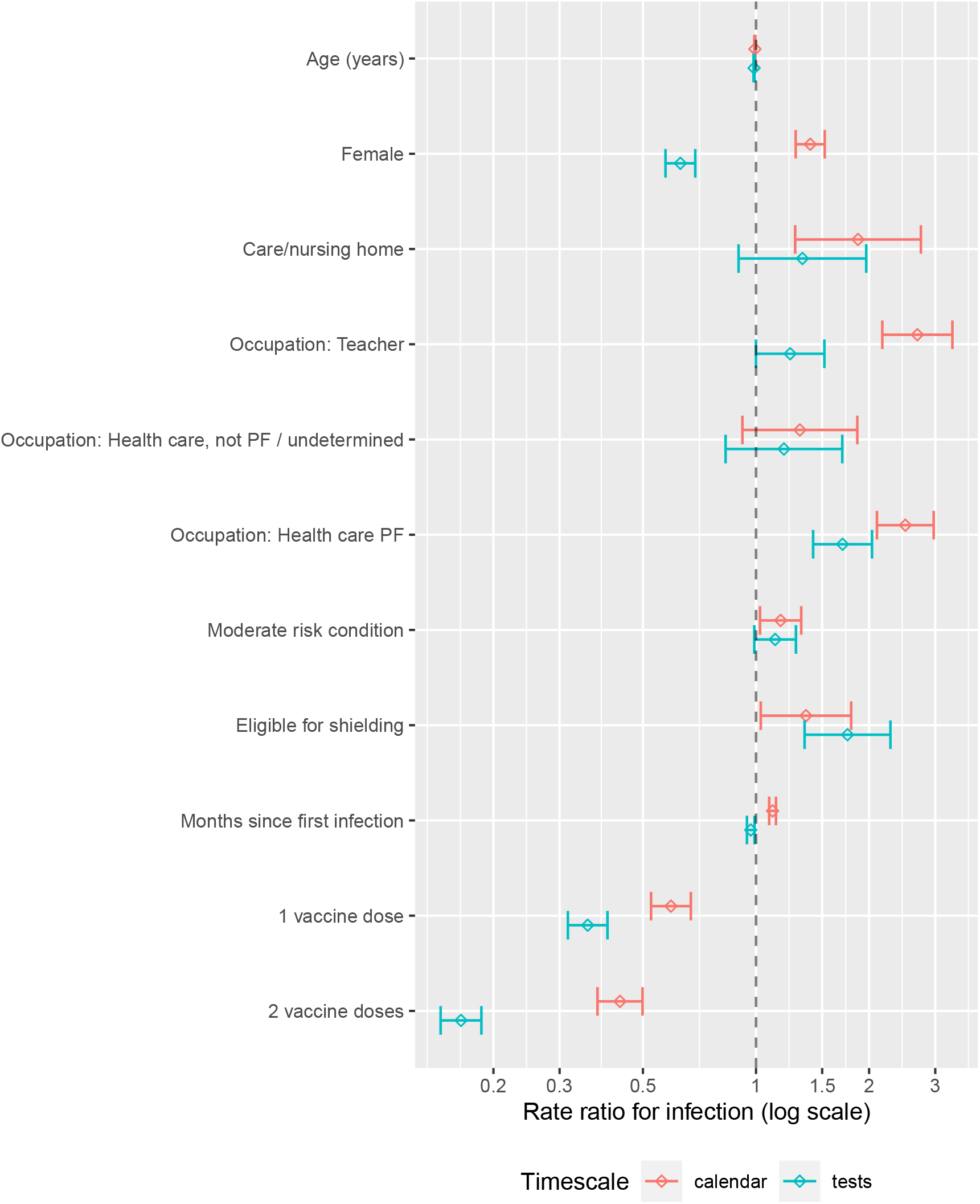
Comparison of Cox regression models for detected reinfection with calendar timescale and tests as timescale. PF, patient-facing.

In the model with calendar timescale, several factors that were associated with higher testing rates – female sex, care home residence, and occupation – were associated with detected reinfection. In the model with tests as timescale these associations were reduced or reversed, consistent with detection bias as the explanation for the effects estimated with calendar timescale. In both models vaccination was associated with lower rates of reinfection. With tests as timescale the efficacy of vaccination against reinfection was estimated as 64% (95 percent CI 60% to 68%) for one dose of vaccine and 84% (95 percent CI 81% to 86%) for two doses. With tests as timescale the estimated efficacy of vaccination was higher (rate ratios were lower) than with calendar timescale, suggesting that the efficacy estimated from the model with calendar timescale may have been biased downwards by lower detection rates in unvaccinated individuals.

Teaching and health care occupations were associated with increased risk of detected reinfection but the rate ratios were lower in the model with tests as timescale than in the model with calendar timescale, suggesting that much of this excess may be driven by increased testing. In both models clinical risk conditions were associated with increased risk of reinfection; a possible explanation for this is that clinical risk conditions are associated with higher exposure to nosocomial transmission [6]. Time since initial infection was associated with higher rates of reinfection in the model with calendar timescale but this association was reversed in the model with tests as timescale.

### Effect of vaccination and other covariates on hospitalised or fatal reinfection

Table 1 shows that hospitalised or fatal reinfection was associated with older age (median age 54 years in cases versus 38 years in noncases) and clinical risk conditions. The numbers of hospitalised cases were too small for associations with occupation to be estimated reliably. The efficacy of two doses of vaccine against hospitalised reinfection was estimated as 71% (95 percent CI 29% to 88%).

**Table 1:** Rate ratios for hospitalised or fatal reinfection in cohort at risk of reinfection

|  | Non-case intervals<br>(1928802) | Cases (60) | Rate ratio (95%<br>CI) | *p*-value |
| --- | --- | --- | --- | --- |
| Age (years) | 38 (IQR 24 to 54) | 55.5 (IQR 34.2 to 72.5) | 1.03 (1.01, 1.04) | 0.004 |
| Female | 1001602 (52%) | 39 (65%) | 1.71 (1.00, 2.92) | 0.05 |
| Care/nursing home | 21178 (1%) | 7 (12%) | 2.98 (1.14, 7.84) | 0.03 |
| **Occupation** |  |  |  |  |
| Other / undetermined | 1800545 (93%) | 57 (95%) | . | . |
| Teacher | 32763 (2%) | 0 (0%) | 0.00 (0.00, Inf) | 1 |
| Health care, not PF /<br>undetermined | 28186 (1%) | 1 (2%) | 1.32 (0.18, 9.67) | 0.8 |
| Health care PF | 67308 (3%) | 2 (3%) | 1.32 (0.31, 5.56) | 0.7 |
| **Clinical risk category** |  |  |  |  |
| No risk condition | 1575010 (82%) | 29 (48%) | . | . |
| Moderate risk condition | 302916 (16%) | 22 (37%) | 2.87 (1.54, 5.38) | $9 \times 10^{-4}$ |
| Eligible for shielding | 50876 (3%) | 9 (15%) | 6.4 (2.8, 14.9) | $1 \times 10^{-5}$ |
| Months since first infection | 5.8 (IQR 4 to 7.7) | 5.1 (IQR 3.6 to 7.3) | 0.95 (0.83, 1.08) | 0.4 |
| **Vaccination status (time-updated)** |  |  |  |  |
| Unvaccinated | 877508 (45%) | 31 (52%) | . | . |
| 1 dose | 464312 (24%) | 14 (23%) | 0.49 (0.24, 1.02) | 0.06 |
| 2 doses | 586982 (30%) | 15 (25%) | 0.29 (0.12, 0.71) | 0.006 |

For age and months since first infection, median and interquartile range (IQR) is given
Vaccination status defined as number of doses received at least 14 days before presentation date.
PF, patient-facing
Rate ratios estimated in a Cox regression model that includes all covariates in the table

## Discussion

### Statement of principal findings

- Among those who are unvaccinated, previous infection with SARS-CoV-2 is associated with a reduction of about 70% in detected new infections and hospitalisations with COVID-19. The protection against new infection is likely to be underestimated because of higher testing rates in those who have previously tested positive, and the protection against severe COVID-19 is likely to be underestimated because specificity (for severe COVID-19) of hospitalisation with COVID-19 is lower in those who have previously tested positive.
- Efficacy of vaccination against reinfection and against hospitalisation with COVID-19 is high in those with previous infection. The combination of natural infection and two doses of vaccine provides maximal protection against new infection with COVID-19: in this group the level of protection is only slightly less in those who were vaccinated at first infection than in those who were unvaccinated at first infection.

### Strengths and limitations

Strengths of this study are the large cohort of test-positive individuals based on ascertainment of all detected infections in the population, the comprehensive linkage to electronic health records, and the ability to examine associations with occupation. Although reinfections with new strains were not confirmed by sequencing, the 90-day interval should be enough to exclude persistent infection except in the immunosuppressed. Restriction to those with definite previous infection – at least two positive tests or Ct < 30 – excludes those who tested positive only once before September 2020, when the Lighthouse labs began reporting Ct values. Stratification by calendar time should eliminate almost all confounding by Alpha and Delta variants, as it took only a few weeks for each of these variants to replace pre-existing strains in Scotland.

The main limitation is that without regular scheduled testing, estimates of association with detected reinfection are subject to ascertainment bias. Because testing rates are lower in unvaccinated than in vaccinated individuals in this cohort, the efficacy of vaccination is likely to be underestimated by a model with calendar timescale. We have attempted to overcome this by comparing two alternative models: a conventional Cox regression with calendar timescale, and a Cox regression with tests as timescale to adjust for differential testing rates. The model with tests as timescale is equivalent to a test-negative case-control design in which tests on each individual are numbered sequentially, and sets of cases and controls matched on test number are analysed as a matched case-control design with conditional logistic regression. Conventional test-negative control designs do not match on test number, and may include arbitrary restrictions on the number of test-negative control observations per individual [9–11]. The calendar timescale model assumes that rate ratios for detected infection are equal to rate ratios for infection on a calendar timescale, even though testing rates vary with vaccination status and other covariates. The tests as timescale model assumes that rate ratios for positive tests on a test timescale are equal to rate ratios for infection on a calendar timescale, even though the probability that an infected individual will seek a test is likely to depend on the severity of symptoms which may vary with vaccination status. This assumption is implicit in conventional test-negative case-control designs also. Even though neither of these two models is likely to be correct, comparison of them allows assessment of the robustness of the estimates to violation of assumptions about how testing rates depend on covariates.

### Relation to other studies

In a recent systematic review of ten cohort studies reported up to 8 July 2021, the average reduction in infection rates in those with previous infection compared with those without evidence of previous infection was 90% [1]. Only three of these studies extended into the period when vaccines became available, and none were able to study a population in which Delta had become the predominant variant. In studies based on case ascertainment through unscheduled testing, the likely size and direction of ascertainment bias cannot be assessed unless testing rates in exposed and unexposed groups are reported. Reinfection rates have been estimated in two UK studies of cohorts in which participants were tested regularly: in the SIREN study of health-care workers, the rate of reinfection up to January 2021 was 2.3 per 1000 person-months [12]; while in the ONS COVID-19 Infection Survey [13], the rate of reinfection up to June 2021 was 4.6 per 1000 person-months. These rates are rather lower than the rate of 6.8 (95% CI 6.4 to 7.2) that we report for unvaccinated individuals: a possible explanation may be that reinfection rates have increased since Delta became the dominant variant in Scotland during May-June 2021.

In previous studies we have used severe COVID-19 – defined by entry to critical care or fatal outcome – as the main outcome measure [4]. As there were few severe cases in the cohort at risk of reinfection we have reported rates for the broader outcome of hospitalised or fatal COVID-19. The reduction of 68% is similar to an earlier estimate based on individuals who had received a single dose of vaccine [14] but as noted earlier this is likely to underestimate the protection against hospitalisations and deaths caused by COVID-19.

Two other studies have reported estimates of vaccine efficacy against detected reinfection: a case-control study in Kentucky reported a 57% reduction [15], and a cohort study of individuals registered with an Israeli healthcare provider reported a 32% reduction but the numbers of cases were small and the confidence interval was wide [16]. Neither study reported or adjusted for differences in testing rates.

### Interpretation and implications for policy

These results support other estimates that the protection against infection and hospitalised COVID-19 conferred by prior infection is of similar magnitude to that obtained with vaccination. The protection against infection is relevant to policies for vaccination certification [17], as recognized by the European Centre for Disease Prevention in guidance that “For individuals that have recovered from a laboratory-confirmed SARS-CoV-2 infection within 180 days prior to travel, it can be considered to ease quarantine and testing requirements.” [18]. The level of protection conferred by the combination of vaccination and natural infection is only slightly less when vaccination precedes first infection; this is relevant to concerns that vaccination might narrow subsequent immune responses through antigenic imprinting [19] arising from a recent report that N antibody levels after infection are lower in those who had received 2 doses of vaccination before infection [20].

## Supporting information

STROBE checklist for cohort study

## Data Availability

The component datasets used here are available via application to the Public Benefits and Privacy Panel for Health and Social Care at https://www.informationgovernance.scot.nhs.uk/pbpphsc/. All source code used for derivation of variables, statistical analysis and generation of this manuscript is available on https://github.com/pmckeigue/covid-scotland_public.

## Declarations

### Public and Patient Involvement statement

This study was conducted under approvals from the Public Benefit and Privacy Panel for Health and Social Care which includes public and patient representatives.

### Ethics approval

This study was performed within Public Health Scotland as part of its statutory duty to monitor and investigate public health problems. Under the [UK Policy Framework for Health and Social Care Research](https://www.hra.nhs.uk/planning-and-improving-research/policies-standards-legislation/uk-policy-framework-health-social-care-research/) set out by the NHS Health Research Authority, this does not fall within the definition of research and ethical review was therefore not required. Individual consent is not required for Public Health Scotland staff to process personal data to perform specific tasks in the public interest that fall within its statutory role. The statutory basis for this is set out in Public Health Scotland’s [privacy notice](https://www.publichealthscotland.scot/our-privacy-notice/personal-data-processing/).

### Transparency declaration

PM as the manuscript’s guarantor affirms: that the manuscript is an honest, accurate, and transparent account of the study being reported; that no important aspects of the study have been omitted; and that any discrepancies from the study as originally planned and registered have been explained. This manuscript has been generated directly from the source data by a reproducible research pipeline.

### Author contributions

Conceptualization: all authors. Formal analysis: PM. Writing original draft: PM, HC. Review and editing: all authors.

## Acknowledgements

We thank Jen Bishop, Bob Taylor and David Caldwell for undertaking the data extraction and linkage.

## Supplementary Figures and Tables

**Figure S1:**
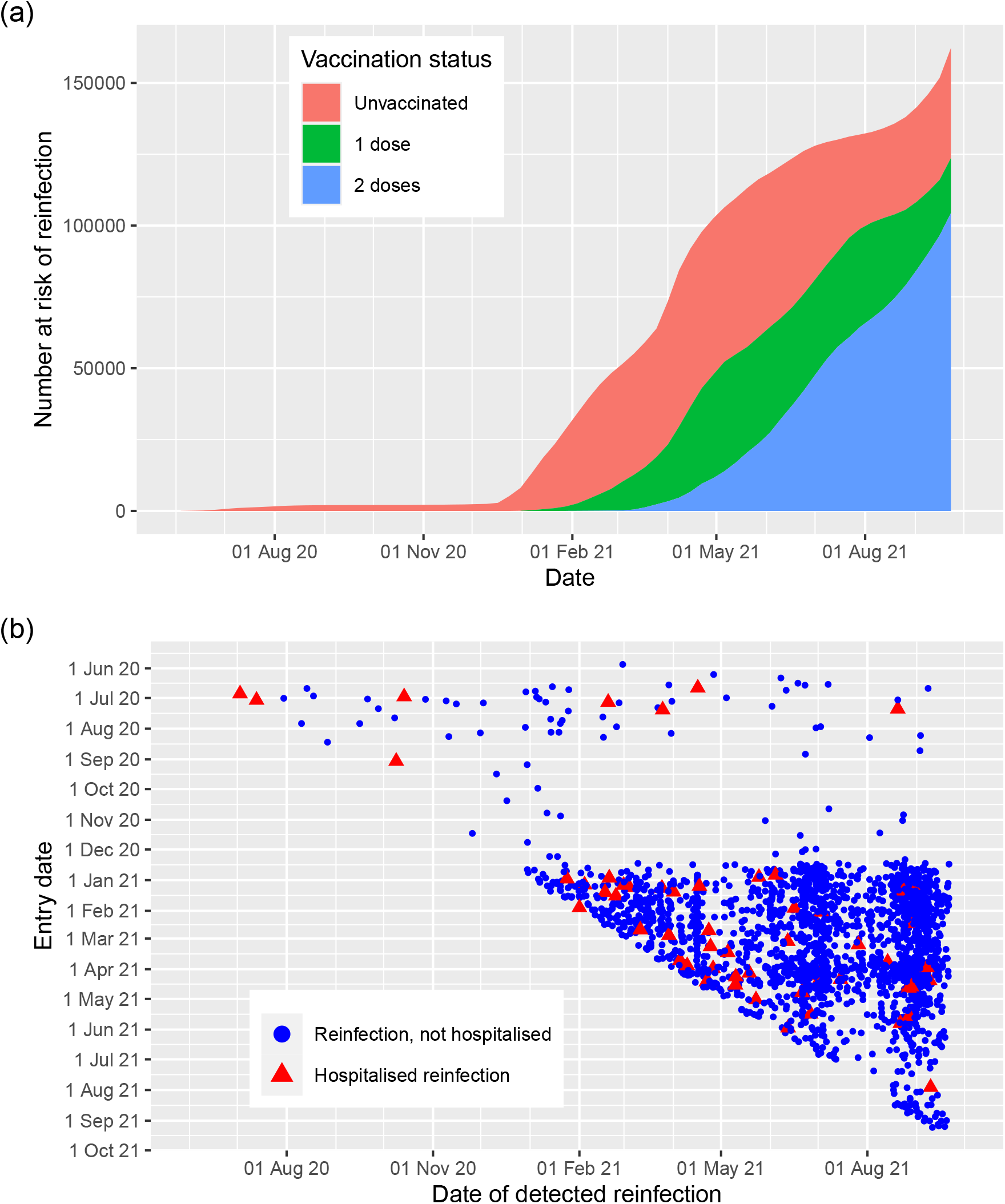
Cohort at risk of reinfection: (a) detected reinfections by entry date and date of positive test; (b) Numbers at risk on each date by vaccination status. Vaccination status is coded as number of doses received at least 14 days before

**Table S1:** Rate of PCR testing for SARS-CoV-2 in cohort at risk for reinfection, by vaccination status and covariates

| Variable | Number of tests |  | Testing rate per person-month |  |
| --- | --- | --- | --- | --- |
|  | 0 or 1 dose | 2 doses | 0 or 1 dose | 2 doses |
| All | 197534 | 201132 | 0.56 | 0.77 |
| **Sex** |  |  |  |  |
| Female | 138098 | 148432 | 0.80 | 0.99 |
| Male | 59436 | 52700 | 0.33 | 0.47 |
| **Age group** |  |  |  |  |
| 0-60 years | 177476 | 163312 | 0.53 | 0.85 |
| 60-74 years | 14235 | 31662 | 1.01 | 0.59 |
| 75+ years | 5823 | 6158 | 0.98 | 0.35 |
| **Residence** |  |  |  |  |
| Independent | 193620 | 197624 | 0.55 | 0.77 |
| Care/nursing home | 3914 | 3508 | 1.50 | 0.71 |
| **Occupation** |  |  |  |  |
| Other / undetermined | 175083 | 174203 | 0.52 | 0.74 |
| Health care PF | 8945 | 15876 | 1.77 | 0.90 |
| Teacher | 10489 | 5224 | 1.96 | 1.21 |
| Health care, not PF / undetermined | 3017 | 5829 | 1.23 | 0.85 |
| **Clinical risk category** |  |  |  |  |
| No risk condition | 164664 | 156385 | 0.53 | 0.81 |
| Moderate risk condition | 28799 | 37832 | 0.78 | 0.65 |
| Eligible for shielding | 4071 | 6915 | 0.93 | 0.57 |

Vaccination status defined as number of doses received at least 14 days before test date.
PF, patient-facing

**Table S2:** Rate ratios for detected reinfection in cohort at risk of reinfection: comparison of models with calendar timescale and tests as timescale

|  | Non-case intervals<br>(1926743) | Cases (2119) | Calendar timescale |  | Tests as timescale |  |
| --- | --- | --- | --- | --- | --- | --- |
|  |  |  | Rate ratio (95%<br>CI) | *p*-value | Rate ratio (95%<br>CI) | *p*-value |
| Age (years) | 38 (IQR 24 to 54) | 31 (IQR 21 to 48) | 0.99 (0.99, 1.00) | $9 \times 10^{-6}$ | 0.99 (0.98, 0.99) | $3 \times 10^{-13}$ |
| Female | 1000346 (52%) | 1295 (61%) | 1.39 (1.28, 1.52) | $3 \times 10^{-13}$ | 0.63 (0.57, 0.69) | $2 \times 10^{-23}$ |
| Care/nursing home | 21154 (1%) | 31 (1%) | 1.87 (1.27, 2.75) | 0.001 | 1.33 (0.90, 1.96) | 0.2 |
| **Occupation** |  |  |  |  |  |  |
| Other / undetermined | 1798753 (93%) | 1849 (87%) | . | . | . | . |
| Teacher | 32674 (2%) | 89 (4%) | 2.69 (2.17, 3.33) | $2 \times 10^{-19}$ | 1.23 (1.00, 1.52) | 0.05 |
| Health care, not PF /<br>undetermined | 28155 (1%) | 32 (2%) | 1.31 (0.92, 1.86) | 0.1 | 1.19 (0.83, 1.70) | 0.3 |
| Health care PF | 67161 (3%) | 149 (7%) | 2.50 (2.10, 2.97) | $6 \times 10^{-25}$ | 1.70 (1.42, 2.04) | $8 \times 10^{-9}$ |
| **Clinical risk category** |  |  |  |  |  |  |
| No risk condition | 1573291 (82%) | 1748 (82%) | . | . | . | . |
| Moderate risk condition | 302622 (16%) | 316 (15%) | 1.16 (1.02, 1.32) | 0.02 | 1.12 (0.99, 1.28) | 0.07 |
| Eligible for shielding | 50830 (3%) | 55 (3%) | 1.36 (1.03, 1.79) | 0.03 | 1.75 (1.35, 2.28) | $3 \times 10^{-5}$ |
| Months since first infection | 5.8 (IQR 4 to 7.7) | 6.9 (IQR 4.8 to 8.6) | 1.11 (1.08, 1.13) | $2 \times 10^{-21}$ | 0.97 (0.95, 0.99) | 0.006 |
| **Vaccination status (time-updated)** |  |  |  |  |  |  |
| Unvaccinated | 876376 (45%) | 1163 (55%) | . | . | . | . |
| 1 dose | 463917 (24%) | 409 (19%) | 0.59 (0.53, 0.67) | $5 \times 10^{-17}$ | 0.36 (0.32, 0.40) | $2 \times 10^{-62}$ |
| 2 doses | 586450 (30%) | 547 (26%) | 0.43 (0.38, 0.50) | $2 \times 10^{-32}$ | 0.16 (0.14, 0.19) | $5 \times 10^{-177}$ |

For each numeric variable, median and interquartile range (IQR) is given.
Vaccination status defined as number of doses received at least 14 days before presentation date.
PF, patient-facing
Rate ratios estimated in Cox regression models that include all covariates in the table
The model with tests as timescale is additionally stratified by 14-day calendar time interval.

